# Inflammatory biomarkers and perinatal depression: a systematic review

**DOI:** 10.1101/2023.01.11.23284231

**Authors:** Anabela Silva-Fernandes, Ana Conde, Margarida Marques, Rafael A. Caparros-Gonzalez, Emma Fransson, Ana Raquel Mesquita, Bárbara Figueiredo, Alkistis Skalkidou

**Affiliations:** Center for Research in Psychology (CIPsi), School of Psychology, University of Minho, Portugal; Portucalense Institute for Human Development. Portucalense University, Portugal; Faculdade de Psicologia, Universidade de Lisboa, Lisbon, Portugal; Faculty of Health Sciences, Department of Nursing, University of Granada, Spain; Instituto de Investigación Biosanitaria ibs.GRANADA, Granada, Spain; Department of Women’s and Children’s Health, Uppsala University, Sweden

**Keywords:** depression, pregnancy, postpartum, perinatal, immune system, biomarkers, cytokines, inflammation

## Abstract

**Background:** Approximately 10 to 20% of pregnant women worldwide experience perinatal depression (PND), a depressive episode with onset during pregnancy or after childbirth. We performed a systematic review to identify, summarize and discuss studies on inflammatory biomarkers described in relation to PND.

**Methods:** Inclusion criteria defined the selection of observational studies written in English, French, Spanish or Portuguese, that evaluate analytical levels of inflammatory molecules (protein levels) in biological fluids in women, with a diagnosis of depression using ICD/DSM diagnostic criteria or depressive symptoms assessed by standardized psychometric instruments, during pregnancy and/or postpartum. Case reports, experimental studies, reviews, qualitative analysis, meta-analysis, gray literature or replicated data were excluded. Three electronic databases were used for search (Pubmed, Web of Science and PsychInfo) and quality assessment of selected studies were performed using the Newcastle-Ottawa Scale. Data extraction included study design; number of subjects; obstetric information; tools and timepoints of depression and inflammatory markers assessment.

**Results:** 56 studies where the major aim was to analyze the association between depression and inflammatory biomarkers during pregnancy and postpartum period were included in this systematic review. Overall, the findings of our systematic review lend support to the hypothesis that several inflammatory markers may be associated with peripartum depressive symptoms. The associations were somewhat different looking at pregnancy compared to the delivery time-point and postpartum, and mainly referred to increased levels of IL-6, IL-8, CRP and TNF-α among depressed.

**Discussion:** Our results revealed high heterogeneity in relation to the timing of biological sampling for markers, as well as timing and instruments used for depression assessment within the perinatal period for the different studies. Studies differed also in relation to use of biomarkers or depression as exposure and outcome respectively, and whether these were addressed at the same timepoint or separate ones.

Given the high burden of PND on women, children and families, it is crucial to try to harmonize methods used in related studies, in order to be able to pool results that could give us insights into the pathophysiological mechanisms behind how the immune system and PND are connected; this could have great impact on early detection, prevention and even treatment of PND.

## 1. Introduction

Pregnancy and postpartum are critical periods for the mental health of the mother, her baby and the whole family. During the past decades, knowledge regarding the psychobiological pathways impacting on mental health has expanded substantially, including studies in the perinatal setting. While epidemiological literature still dominates the perinatal mental health field, increasing evidence supports the link between psychosocial and biological pathways, and more studies focus on the role of the immune system in the development of perinatal depression (PND), both with antenatal (AND) and postpartum onset (PPD) (e.g., (1-4)).

The immune system is a complex network that aims to protect the host from invading microorganisms and induce wound healing when needed. During an immune response, such as the system’s reaction to an infection, human behavior is affected, leading often to increased inactivity, increased sleepiness, decreased appetite, and social withdrawal, behaviors that also resemble those characteristic of clinical depression (5, 6). These so-called sickness behaviors can also be induced by cytokines or experimental exposure to endotoxins (7, 8).

The interplay between the immune functioning and depression has been explored over the last decades and a bidirectional loop has been described. While inflammation seems to play a key role in depression’s pathogenesis, at least for a subset of depressed individuals, it was also been shown that depression, adversity and stress have also been associated to exaggerated or prolonged inflammatory responses (for a review see (9). Thus, studying the pathways involved in this interplay could allow to identify inflammatory biomarkers with the potential to predict depression, but also possible targets for prevention and/or treatment.

As many bodily systems, the immune system undergoes large changes to adapt to the pregnancy and postpartum period. An inflammatory response is necessary to promote implantation (10, 11), while during pregnancy, the maternal immune system is challenged by the semi-allogeneic fetus and needs to adapt, mainly by a shift towards immune tolerance, characterized i.e. by an increase in regulatory T-cells (Tregs) as well as macrophages type 2 (M2) (12, 13).

The dramatic shift in the characteristics of the immune response during pregnancy and postpartum seem to impact on maternal pregnancy mood and in some way differently across time (14). The increase in Treg occurring in mid-pregnancy coincide with the pregnancy period when most women report increased wellbeing, and Treg activity has been associated with resilience to stress in animal studies (15). Furthermore, women with chronic autoimmune conditions, such as multiple sclerosis (MS) and rheumatoid arthritis (RA) often experience remission of the conditions during this pregnancy period (16). In the third trimester, pro-inflammatory activity rise and the delivery itself could be characterized as a largely pro-inflammatory event, including the remodeling of the cervical tissue as well as uterine contractions (17, 18). This intensification of pro-inflammatory activity in late pregnancy co-occurs with an increase in depressive symptoms during this period, compared to lower levels in the second trimester (19).

After childbirth, maternal body needs to reduce the pro-inflammatory activity, and a decrease of many inflammatory markers has been reported from the third trimester to the postpartum (20, 21). For many women, this period is characterized by additional bodily changes associated with wound healing and breast-feeding onset that could also impact on immune actions. Furthermore, sleep disturbances that are common perinatally, are also known to induce a pro-inflammatory state (22) affecting depression risk. The early postpartum period has been characterized by a drop in Treg cells, and immune response characteristic of T helper 1 (Th1) or Macrophages type 1 has been described (23). While MS and RA often improves during pregnancy, these conditions worsen again postpartum (16). These changes in the immune system seems also to be associated to an increased risk of depression during the postpartum period (e.g., (1, 24, 25)).

A previous systematic review on postpartum depression included 25 articles (26). Their most robust finding was that levels of CRP in late pregnancy could predict postpartum depression. However, many new studies on this topic have been published and techniques for biomarker analyses have developed further and more focus has been given to depression with antenatal onset. A recent meta-analyses of inflammatory markers for major depression in general and specifically perinatally, showed robust results of increase of several pro-inflammatory markers in depressed individuals (27). Some studies of specific inflammatory biomarkers in the perinatal setting show conflicting results. A more recent review of literature has precisely reflected on the distinct and changing inflammatory profiles throughout pregnancy and postpartum which makes the study of depression-related alterations in these periods highly complex (14). Contributing to this complexity is the fact that many studies in the field utilize diverse methodologies that make difficult to compare findings in order to understand the interplay between the biological processes involving the adjustments of the immune system during pregnancy and postpartum and the trajectories of change of mental health during this period. Despite the growing evidence in the field of immune related biomarkers in PND and PPD, clinical applications for biomarkers for depression prediction or treatment during these periods are lacking in clinical practice.

In order to address this shortcoming of earlier studies and achieve a better categorization of the scientific output exploring the associations between the functioning of the maternal immune system and PND, we performed a systematic review involving all the original empirical quantitative studies conducted during the perinatal period, which assessed both levels of women’s inflammatory molecules as well as depression (diagnosis of depression or depressive symptoms or depressive symptomatology), while (1) taking into account whether the assessments of the two variables were performed cross-sectionally at one time-point or longitudinally and even (2) if one of the variables could be used as a predictor for the other.

## 2. Methods

This study followed the recommendations outlined in the Preferred Reporting Items for Systematic Review and Meta-Analysis (PRISMA) guidelines (28, 29) and has been registered on PROSPERO (Registration ID: CRD42020210080). Protocol details are available at https://www.crd.york.ac.uk/PROSPERO/display_record.php?RecordID=210080.

### 2.1. Search strategy

An initial article search was conducted at July 3^rd^, 2020, through three electronic databases (Pubmed, Web of Science and PsychInfo) to identify English, Portuguese, French and Spanish-language human studies unrestricted by year of publication (for search terms see S1 Table). In February 23^rd^, 2022, a new search was conducted using the same search formula and filters (with the exception of the Filter “Journal Article” in Pubmed that is no longer available) to determine new entries. After duplicates removal, the unique entries from this new search were identified by table comparison.

Duplicates detection was performed by two independent reviewers (ASF and MM) on two different platforms (Endnote and Rayyan) using manual review by ordering articles by title, authors, pages, journal. Finally, the authors met to reach accordance on the final number of duplicates.

### 2.2. Studies selection

Original quantitative studies that evaluate the levels of inflammatory molecules in women with a diagnosis of depression or depressive symptomatology conducted in women during pregnancy and/or at the postpartum period were eligible for this systematic review. Two authors (ASF and MM) screened the titles and abstracts of articles from the primary search independently against inclusion and exclusion criteria:

Inclusion criteria:

- Written in English or French or Spanish or Portuguese
- Observational studies
- Depression assessed using ICD/DSM diagnostic criteria either through diagnostic interview or expert opinion. Alternatively, depressive symptomatology assessed using standardized psychometric instruments
- Inflammatory molecules measurement (protein levels) in biologic fluids using analytical techniques

Exclusion criteria:

- Case reports or experimental studies or reviews or qualitative analysis or meta-analysis or gray literature
- Replicated data

The full text of qualifying articles was then assessed against the same standard by different pairs of authors (ASF&MS; AC&ARM; BF&RCG; AS&MM). Any discrepancies were resolved first through discussion amongst the pair, and if a consensus could still not be reached, by conferring with other group members.

### 2.3 Data extraction

The following data was extracted from each selected study: country of origin; study design; number of subjects; socio-economic status/ethnicity; obstetric information, namely delivery mode; assessment of depression: instrument(s) and timepoint(s) and inflammatory protein markers: biological fluid/hour of collection, timepoint(s), dosage assessment technique, inflammatory markers and results.

### 2.4 Quality assessment

Following PRISMA guidelines, the quality assessment of selected studies (RCG&MM) and the data extraction were conducted independently by two authors. At the end, the complete data extraction table was revised to uniformization. Different version of the Newcastle-Ottawa Scale (NOS) were used to assess the methodological quality of selected studies (30-32). All the inter-rater agreements between authors were verified prior to resolving disagreements.

## 3. Results

A total of 3527 relevant references were initially identified in an electronic search of three databases: Pubmed, Web of Science and PsychInfo. All 806 duplicated references were removed, and 2721 articles remained. The titles and abstracts of the identified references were screened, and 2594 non-relevant references were excluded. The full text of the 127 remaining studies was then screened, and 44 studies met one or more exclusion criteria. At the final stage, 83 studies were included in the review. A flow diagram of the search selection for the included studies is presented in Fig. 1.

**Fig 1.**
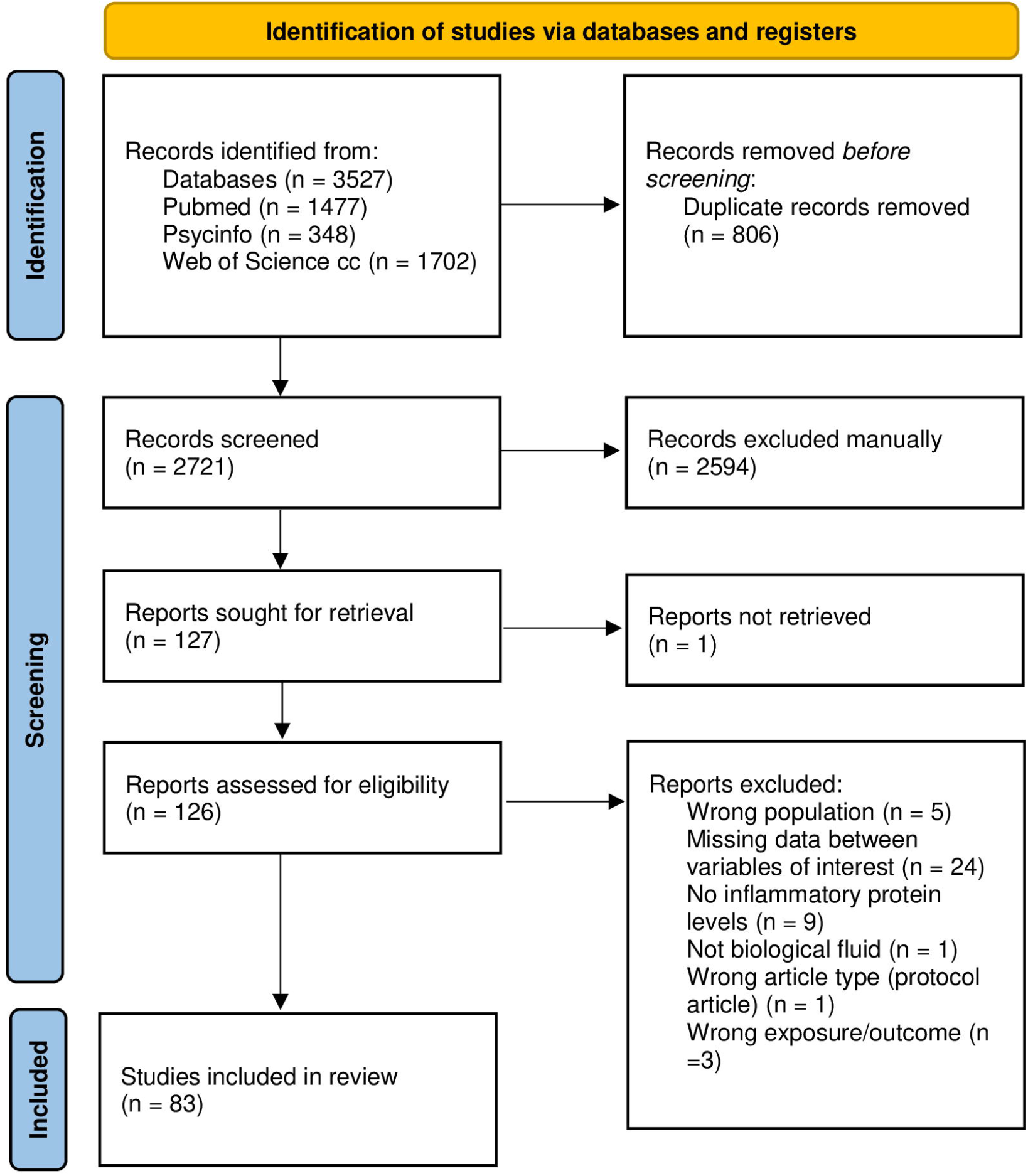
The PRISMA flow diagram. *Consider, if feasible to do so, reporting the number of records identified from each database or register searched (rather than the total number across all databases/registers). **If automation tools were used, indicate how many records were excluded by a human and how many were excluded by automation tools. *From:* Page MJ, McKenzie JE, Bossuyt PM, Boutron I, Hoffmann TC, Mulrow CD, et al. The PRISMA 2020 statement: an updated guideline for reporting systematic reviews. BMJ 2021;372:n71. doi: 10.1136/bmj.n71 For more information, visit: http://www.prisma-statement.org/

From the 83 studies included in the qualitative analysis 27 were studies in which the association between depression and inflammatory markers wasn’t the principal aim but since this derived data was available, we performed data extraction from these studies and presented them in the S2 Table.

For the accomplishment of the aims of this systematic review we have focused the 56 studies where the major aim was to analyze the association between depression and inflammatory biomarkers during pregnancy and the postpartum period. Considering the study design used, 8 studies (3 of which with prospective analysis) used a case-control design, 20 studies a cross-sectional, and 27 a cohort analysis design. Thirty one studies involved repeated assessment time points and longitudinal analyses both during pregnancy (4 studies) and from pregnancy to postpartum (27 studies).

The studies included originate from different countries and research settings, while the involved participants present with markedly different sociodemographic and clinical features (e.g., with and without psychosocial risks). Further, depression was assessed with different methods, from self-reported questionnaires for assessing depressive symptoms, depressive mood, depressive symptomatology or clinical interviews for diagnosing depressive disorders. Lastly, inflammation-related molecules were assessed with very different techniques.

Due to this diversity, results are presented in two steps. Firstly, cross-sectional, and case-control studies on inflammatory biomarkers and depression are presented accordingly to the period involved (pregnancy or postpartum) (for details see Table 1). Secondly, longitudinal studies are presented, with a note on whether a study included predictive analyses (for details see Table 2).

**Table 1.** Cross-sectional and case-control studies on inflammatory biomarkers and depression

| Reference | Country | Study design | Number of subjects | Socio-economic status/ethnicity | Delivery mode | Assesment of depression |  | Inflammatory protein markers |  |  |  | Results | Total quality score |
| --- | --- | --- | --- | --- | --- | --- | --- | --- | --- | --- | --- | --- | --- |
|  |  |  |  |  |  | Instruments | Timepoint(s) | Biological fluid/hour of collection | Timepoint(s) | Dosage assesment technique | Inflammatory markers |  |  |
| Pregnancy |  |  |  |  |  |  |  |  |  |  |  |  |  |
| (34) | USA | Nested case-control (from a prospectiv e cohort study) | T=462 (Ct=390, PND=72) | 39.5% college graduate; 40.7% non-white race; | NR | Perinatal depression documented by the obstetric or the primary care provider in the electronic medical record (EMR) of the study institution. | Anytime across pregnancy | Plasma/NR | Average of three visits: 10, 18 and 26 gw | (1) Hs Multiplex Assay; (2) DuoSet ELISA | (1) IL-6, IL-10, CRP, IL-1β, TNF-α, (2) CRP | NS | 6 |
| (42) | USA | Cross-sectional | T=187 | 87.2% ≥High school education; 87.7% reported income of \$34,315.2±31,431.2; 100% African American | NR | Depressive symptoms measured as CES-D total score; CES-D ≥ 16 equate with symptoms of depression; | 13.1-28.6 gw | Serum/NR | 13.1-28.6 gw | (1) ELISA; (2) custom 4-plex assay | (1) hs-CRP, (2) IL-1β, IL-6, IL-10 and TNF-α | (+) correlation of CES-D with IL-6 (p=.04) and IL-1β (p=.03) | 7 |
| (43) | Finland | Cross-sectional (from a cohort study) | T=139 | 5.8% low education levels | NR | EPDS: continuous total sum score was used for the main analyses. Additional comparisons (high/low EPDS) with the cut-point 9/10. | 24 gw | Serum/randomly through the office hours | 24 gw | Bio-Plex Pro Human Cytokine 21- and 27-Plex Assay kits | IL-6, IL-10, TNF-α, IL-9, IL-5, IL-12, IL-13, IFN-γ, IL-4 | (+) correlations between EPDS scores and four cytokines: IL-5 (p=.007), IL-9 (p=.011), IL-12 (p=.018), IL-13 (p=.029) and the IFN-γ/IL-4 ratio (p=.039). | 6 |
| (45) | USA | Cross-sectional | T=60 | 82% high school or less education; 63% annual family income < \$15,000; 57% African American | NR | Depressive symptoms measured as CES-D total score (clinical cut-off 16) | M=15±7.8 gw | Serum/9:30 am-1:30 pm | 15±7.8 gw | Ultra sensitive multiplex kits | IL-6, TNF-α | NS | 7 |
| (35) | USA | Nested case-control (from a cohort study) | T=200 (Ct=100; depressed =100) | Caucasian: 81% in depressed group and 78% in Cts | NR | DSM-V diagnosis of depression (depressed group). Documented negative EPDS for Cts | Prior to pregnancy and confirmed during pregnancy | Serum/NR | 11-14 gw (M=12.8 gw in Cts and 12.6 gw in depressed group) | (1) hs-ELISA; (2) Multiplex Assay | (1) IL-6, (2) TNF-α | ↑ IL-6 (p=.03) and ↑ TNF-α (p<.0001) levels in depressed vs Cts groups. | 6 |
| (46) | Thailand | Cross sectional | T=73 (Non-pregnant= 24; pregnant no PND=25, pregnant + PND=24) | Years of education: 17.8±1.3 Non-pregnant; 12.5±3.3 pregnant no PND; 12.0±4.3 pregnant + PND. | NR | EPDS to asses the diagnosis of perinatal depression (EPDS≥11 for depressed and EPDS≤2 for non-depressed) and HAM-D | 3rd trimester (at the end of term) and 4-6 wks PP | Serum | 3rd trimester (at the end of term) | (1) ELISA; (2) Atomic absorption spectrometric method; (3) latex immunoassay; (4) spectrophotometric method; | (1) assay of IgM/IgA responses to tryptophan + TRYCATS: Anthranilic acid, Xanthurenic acid, Quinolinic acid, Kynurenic acid, IgA Picolinic acid, 3-OH Kynurenine, Kynurenine, Quinaldic acid, | ↓ IgA responses to anthranilic acid in PND vs pregnant no PND (p=.047). | 8 |
|  |  |  |  |  |  |  |  |  |  |  | 3-OH Anthranilic acid; (2) zinc, (3) hs-CRP, and (4) haptoglobin |  |  |
| (36) | Sweden | Case-control (substudy in the BASIC cohort study) | T=258 (Ct=160; Perinatal depressive disorder=59; Antidepressant treatment=39) | University education: Ct=84.4%, perinatal depressive disorder=67.8%, antidepressant treatment=74.4% | NR | (1) EPDS (psychophysiology sub-study EPDS≥13, cesarean EPDS≥17). (2) MINI and MADRS performed only in participants from the psychophysiology sub-study. | (1) 17 and 32 gw; (2) 35-39 gw | Plasma/9:30 am-1:00pm and morning before cesarean | Late pregnancy (35-39 gw) (n=205) and at morning before cesarean section (38w) (n=53) | Proseek Multiplex Inflammation I (based on proximity extension assay technology) | 92 inflammatory proteins | ↓ 23 inflammatory proteins in women with perinatal depression ($p<.01$) and women on SSRI treatment in comparison with Ct ($p<.01$): TRAIL, CSF1, CX3CL1, CST5, DNER, VEGFA, STAMPB, CD5, CD244, TNFRSF9, TNFB, IL10RB, CD40, IL15RA, hGDNF, ST1A1, CCL11, ADA, CCL25, uPA, AXIN1, SLAMF1, IL17C (-) significant correlations between the 23 molecules described above with depression symptoms severity (MADRS scores) at 36 gw as well as with EPDS scores in 17 and 32 gw (with the exception of AXIN1 and TNFRSF9 at 32 gw). | 6 |
| (37) | USA | Cross-sectional (from a prospective observational study) | T=117 | NR | 100% cesarean | IDS-SR30≥18, MINI | Pre-cesarean | Plasma and CSF/NR | Pre-cesarean | Flow cytometry | IL-1β, IFN-α, IFN-γ, TNF-α, MCP-1, IL-6, IL-8, IL-10, IL-12p70, IL-17A, IL-18, IL-23, and IL-33. | ↑ IL-1β (95%CI 5.9-9148.5), ↑ IL-23 (95%CI, 1.7-294.5), ↑ IL-33 (95%CI 1.1-2.6) levels in the CSF have significant associations with increased odds of PND. | - |
| (38) | China (Taiwan) | Case-control | T=33 (Ct=16; PND=17) | Education years: Ct=16.8±1.0, PND=16.6±1.5 | NR | DSM-IV diagnosis by MINI; EPDS≥12/13 | 16-28 gw | Plasma/9:00-10:00am | 16-28 gw | (1) nephelometry immunoassay; (2) Multiplex assay | (1) CRP; (2) IL-6, IL-10 and TNF-α | ↑ TNF-α levels in PND vs Ct groups ($p=.016$). (+) correlation between PND duration and TNF-α levels ($p<.01$) | 6 |
| (33) | USA | Cross-sectional | T=105 | 51.5% High school or less; 4.8% household income <\$4,999; 70.5% caucasian | NR | POMS-D continuous total sum score (7.6% met the protocol's screening criteria for possible clinical depression with a POMS-D score greater than 20) | 16-26 gw (M=20 gw) | Plasma/mean time of day=11:25am | 16-26 gw (M=20 gw) | 12-plex assay | TNF-α, IFN-γ, IL-1β, IL-2, IL-4, IL-5, IL-6, IL-7, IL-8, IL-10, IL-12, and IL-13 | (-) correlations of POMS-D with IL-1β ($p<.05$), IL-7 ($p<.05$) and TNF-α ($p<.01$). Hierarchical linear regression models: depression contributed to the levels of plasma cytokine IL-1β, IL-7 and TNF-α. | 6 |
| (44) | USA | Cross-sectional | T=206 | 95.1% High school or less; 49.5% Mexican American | NR | Depressive symptoms defined as a CES-D | 22-24 gw | Plasma/NR | 22-24 gw | ELISA | IL-1RA | ↑ IL-1RA levels in group with CES-D≥20 in | 8 |
|  |  | (from a prospective observational study) |  |  |  | score of >20. |  |  |  |  |  | comparison with CES-D<20 group (p=.018). |  |
| (91) | USA | Cross-sectional (from a cohort study) | T=72 | African-american; 60% unemployed; 49% high school level or less | NR | CES-D continuous total sum score (CES-D ≥ 16 frequently used to indicate a positive screen for depression) | 14-17 gw | Plasma/NR | 14-17 gw | BioPlex Pro 17-plex kit. | IL-1β, TNF-α, IL-6, IL-8, IL-12, IL-17, IFN-γ | (+) correlations between CES-D scores and IL-8 (p=.008). | 6 |
| (40) | NR | Cross-sectional | T= 44 (Ct=14; PMMD=30) | NR | NR | PMMD monitored by consultation psychiatric service. The severity of depressive symptoms was measured by the EPDS. | M= 32.9 ± 4.18 gw | Plasma/ Late Morning | M= 32.9; SD=4.18 gw | ELISA kit | ERVWE1 | NS | 7 |
| (39) | Italy | Cross-sectional study | T= 79 women (20 with PND+ TRAUMA, ii) 19 with PND + no-TRAUMA, iii) 20 HV pregnant; iv) 20 HV non-pregnant | Years of Education: No trauma=13.0±3.3; Trauma=14.5±2.9; HVpregnancy=14.0±3.9); Unemployed: No trauma= 10.5%; Trauma =25%; HVpregnancy=68.8% | NR | Diagnosis of PND was made by clinical interview according to the DSM-V criteria. The severity of current depressive symptoms was assessed by EPDS (EPDS≥12 for clinically relevant PND) | 22-24 gw | Serum/8:00 and 10:00 a.m | 22-24 gw | (1) NR; (2) ELISA | 1) CRP; 2) TNF-α and IL-6 | ↑ CRP in the PND + no-TRAUMA group compared to the HV pregnant group; | 6 |
| **Postpartum** |  |  |  |  |  |  |  |  |  |  |  |  |  |
| (57) | USA | Cross-sectional (from a longitudinal study) | T=69 | Annual household income <\$15,000: 37.5% African American group and 18.9% in White group. High school graduate or less: 15.6% African American group and 27.0% in White group. African American (n = 32) and White (n = 37) | NR | CES-D continuous total sum score | 7-10 wks PP | Serum and LPS-stimulated PBMCs culture/NR | 7-10 wks PP | (1) single spot ultra-sensitive ELISA; (2) multiplex assay | <u>Serum</u>: (1) IL-6; (2) TNF-α and IL-8<br><u>LPS-stimulated PBMCs</u>: (2) IL-6, TNF-α, IL-8 and IL-1β | Among African American women, controlling for BMI, (+) correlation between stimulated IL-6 (p≤.01), IL-8 (p≤.01) and TNF-α (p=.02) and CES-D depressed mood subscale. | 8 |
| (55) | USA | Cross-sectional (from a RCT study) | T=56 (n=33 for hsIL-6 ELISA) | 23.2% lower SE class; 92.9% Caucasian | NR | (1) HRSD≥15 and (2) clinical interview including SCID-IV (if HRSD≥15 twice within the following 7-day period to identify a depression recurrence). | (1) 2,3,4,6,8,11,14 and 17 wks PP; | Plasma/NR | 2, 3, or 4 wks PP | hsELISA | hsIL-6 | NS (for risk depression recurrence) | 7 |
| (58) | China | Cross-sectional | T=10 (Ct=8; PPD=2) | NR | NR | patients with PPD provided by medical patterns | within the 4 weeks period of giving birth | Plasma/NR | within the 4 weeks period of giving birth | Ultrasensitive Lab-on-chips (LOC) device and ELISA | IL-2, IL-6, IL-8 and TNF-α | ↑ IL-2 and ↑ IL-8 levels in PPD compared to Ct group (no statistical information is reported) | 2 |
| (63) | USA | Cross-sectional | T=181 | Income <\$10,000: 13.6% breastfeeders group and 39.4% formula feeders group. White race: 89.8% breastfeeders group and 73.4% formula feeders group. | NR | POMS-D continuous total sum score | between 4-6 wks PP (M=5.2 wks) | Serum/8:00-11:00 am | between 4-6 wks PP (M=5.2 wks) | ELISA | IFN-γ and IL-10 | (-) correlation between POMS-D and INF-γ only in the formula feeders (p<.05). | 8 |
| (64) | USA | Cross-sectional (secondary analysis) | T=199 (depressed n=25) | Depressed mothers had lower income in comparison with no-depressed women (p<.06) | NR | POMS-D ≥21 (depressed mothers were categorized as those with scores in the highest decile on the POMS-D scale) | 4-6 wks PP (M=5.3 wks PP) | Serum and ex-vivo whole blood culture supernatant/ 8:00-11:00 am | 4-6 wks PP (M=5.3 wks PP) | ELISA | IFN-γ, IL-6 IL-10 | ↓ serum IFN-γ (p<.001), ↓IFN-γ/IL-10 ratio in both serum (p<.04) and whole blood culture supernatants (p<.009) in depressed in comparison with not-depressed. | 6 |
| (62) | Iran | Case-control | T=62 (Cts=30; MDD=10; minor depression =22) | NR | NR | a psychiatrist used DSM-IV-TR to diagnose major and minor depression. | NR | Serum/NR | immediately after parturition | single radial immunodiffusion | IgG, IgM and IgA and complements C3 and C4 | ↑IgG levels in MDD (p=.026) compared with minor depression | 6 |
| (52) | USA | Cross-sectional | T=165 (Cts=60; depressive episode with peripartum onset=87; 18 displayed depression but did not fulfill the time criteria for peripartum-onset depression defined by DSM-5). | Unemployed: Cts=38%; Depressive episode with peripartum onset=37% | NR | EPDS total score; clinical interview SCID-5 for diagnosis of depressive episode with peripartum onset | 8 wks PP (6-12 wks PP) | Plasma/9:00 am-12:00 pm | 8 wks PP (6-12 wks PP) | (1) MesoScale Discovery platform; (2) HPLC; (3) GC-MS; (4) UPLC-MS/MS | (1) IL-1β, IL-2, IL-6, IL-8, IL-10, and TNF-α; (2) Tryptophan, Kynurenine; (3) quinolinic acid; (4) serotonin, kynurenine acid and nicotinic acid | ↑IL-6, ↑IL-8 and ↓serotonin, ↓IL-2 and ↓quinolinic acid were associated with ↑ risk of PPD (OR<sub>IL-6</sub>=3.0, p=.007; OR<sub>IL-8</sub>=3.32, p=.009, per pg/ml increase; OR<sub>serotonin</sub>=1.43, p=.016, per nM decrease; OR<sub>IL-2</sub>=2.34, p=.002, per pg/ml decrease; OR<sub>quinolinic acid</sub>=4.48, p=.014, per nM decrease) and ↑ depressive symptoms (IL-6: p=.022; IL-8: p=.006; serotonin: p=.003; IL-2: p=.005; quinolinic acid: p=.022) ↑kynurenine/serotonin ratio was associated with an increased risk for PPD (OR = 1.35 per unit increase, p=.038) and ↓serotonin/kynurenine ratio was associated with ↑EPDS score (p=.009). | 7 |
| (61) | Japan | Cross-sectional | T=129 breastfeeding mothers | 96.1% high social support; 9 or less years of education: 1.6% | NR | EPDS ≥9 as having postpartum depression. | 3 mo PP | Breast milk/65.9% at 12:00-2:00 pm | 3 mo PP | ELISA | TGF-β2 | ↑TGF-β2 in mothers with depression than mothers without depression (p=.02); depressed mothers had a 3.11-fold (95% CI: 1.03–9.37) higher likelihood to have ↑TGF-β2 levels in their breast milk. | 8 |
| (59) | Sweden | Cross-sectional | T=64 (preterm group=27; | Low annual income (<115,000 SEK); preterm=0%, | NR | Semi-structured interview. Descriptions of | Within 5 days after delivery | Serum/NR | During labor | Multiplex assay | IL-1α, IL-1β, IL-2, IL-4, IL-6, IL-8, IL-10, IL-12 | Pre-term group: (+) correlation between IL-8 and depressive | 6 |
| | | | term group=37) | term=10.8%. Secondary school: preterm=51.8%, term=40.5%. | | continuous depressed mood were considered as depressive symptoms. | | | | | p70, IL-13, IL-17, IL-18 and IFN-γ | symptoms (p<.01) | |
| (56) | USA | Cross-sectional | T=119 | Income mean \$18,000 ± \$6,000. 84% Caucasian | NR | POMS-D total score (cutoff score of 25 above which triggered a referral to a mental health professional.) | 4-6 wks PP (M=4.5 ± 2.3) | Morning hindmilk/pick up visit before 11:30 am | 4-6 wks PP (M=4.5 ± 2.3) | ELISA | IgA | NS | 9 |
| (60) | USA | Cross-sectional (from multicenter urban birth cohort study) | T=469 | 40% less than high school education. 70% annual income <\$15,000. 73% African American | NR | EPDS total score (EPDS≥12 indicated a need for further mental health evaluation) | 89% at 12 mo PP, 8% at 24 mo PP and 3% at 36 mo PP | PBMC cells culture ex-vivo stimulated/NR | 89% at 12 mo PP, 8% at 24 mo PP and 3% at 36 mo PP | Multiplex ELISA assay | Innate Stimuli: IFN-α, IFN-γ, IL-10, IL-12p40, TNF-α and IL-8. Adaptive and Mitogenic Stimuli: IFN-γ, IL-10, IL-13, IL-4 and IL-5 | (-) correlations between depression and innate immune responses, namely, CpG-induced TNF-α (p=.02), RSV induced IL-8 (p<.05), and LPS induced IFN-γ (p=.02). (-) correlations between depression and adaptive immune responses, namely, several DM-induced cytokines (IL-4, IL-5, IL-10, and IL-13; p<.05), and CR-induced IL-10 (p=.01). | 7 |

**Table 2.**
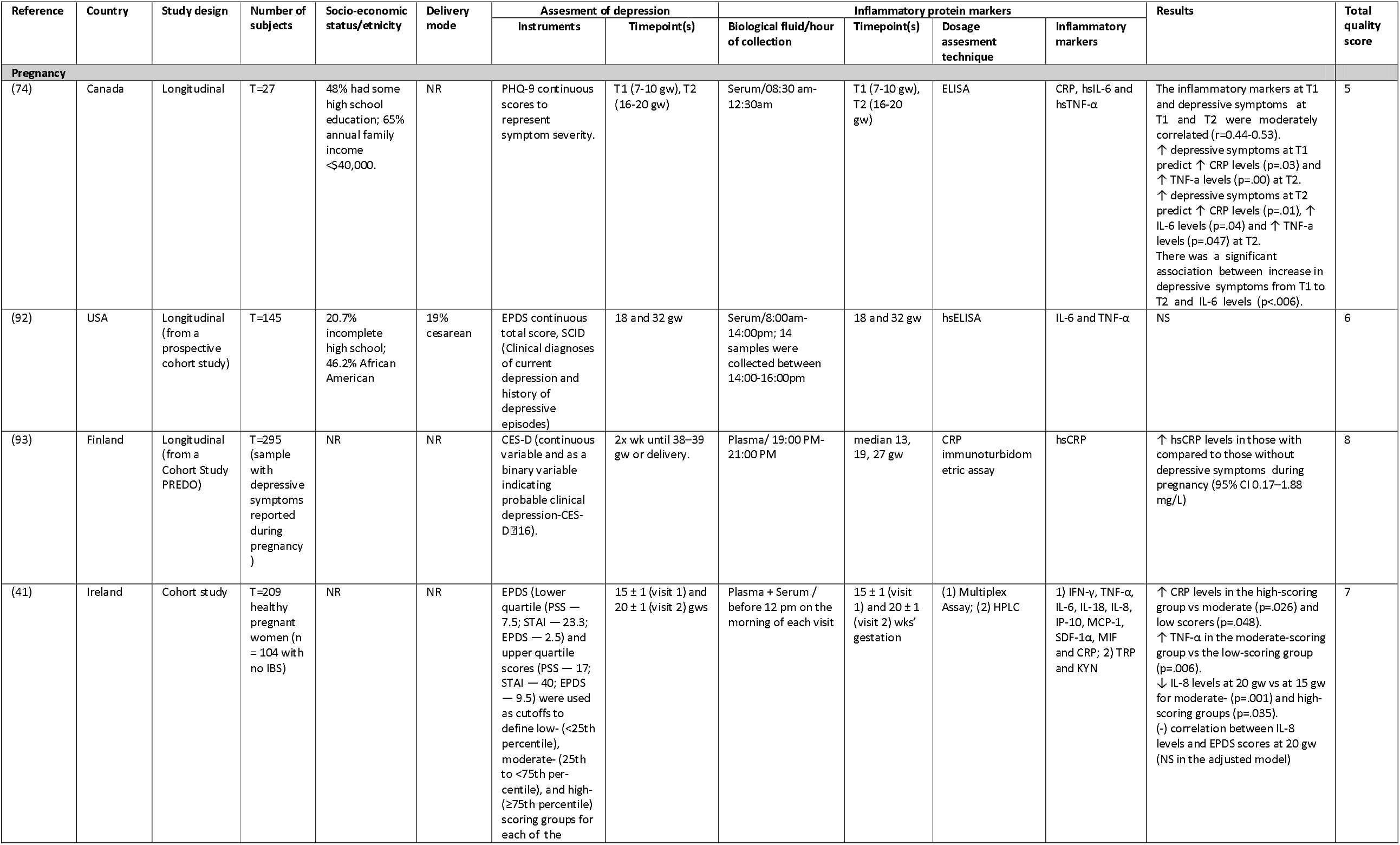
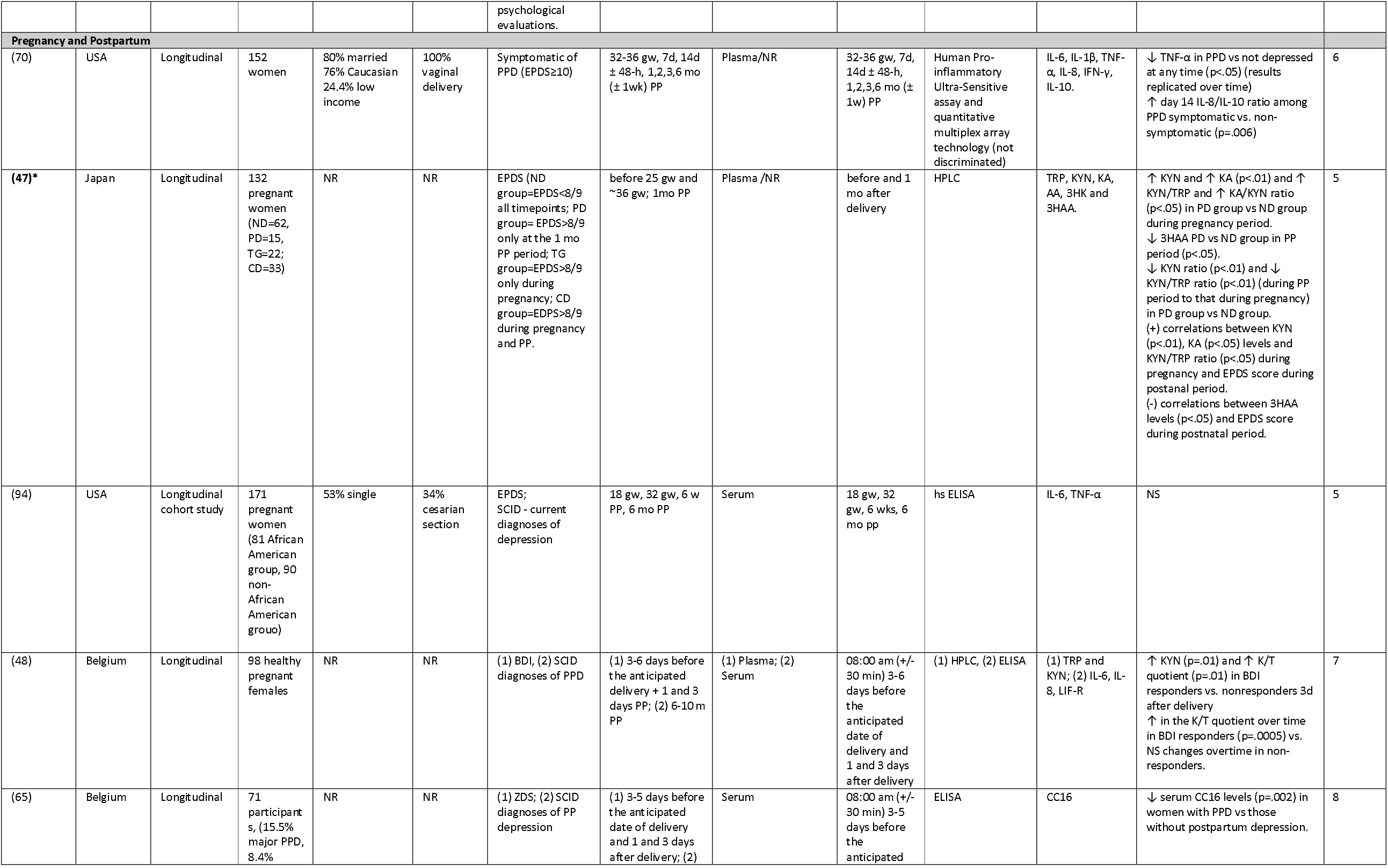
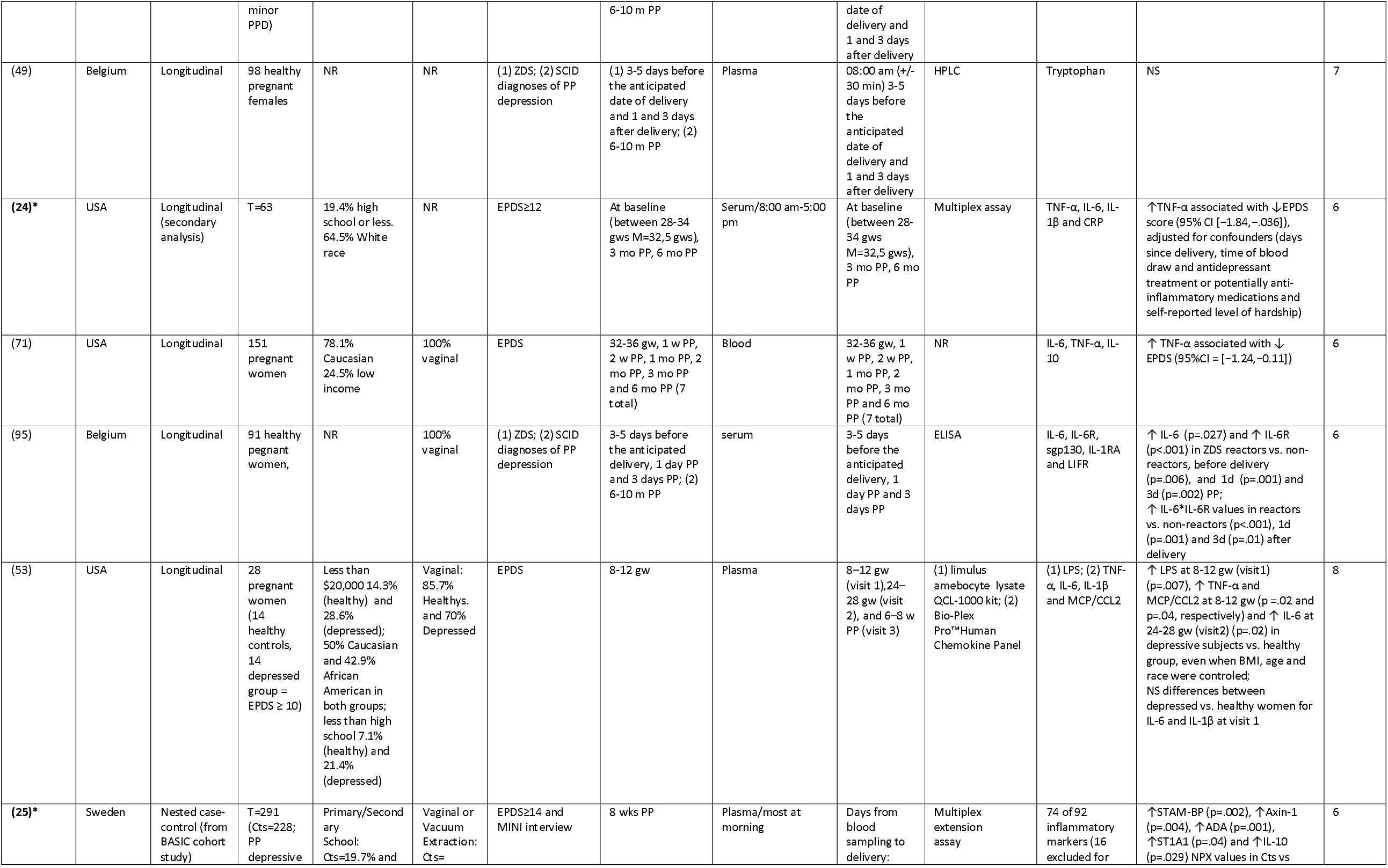
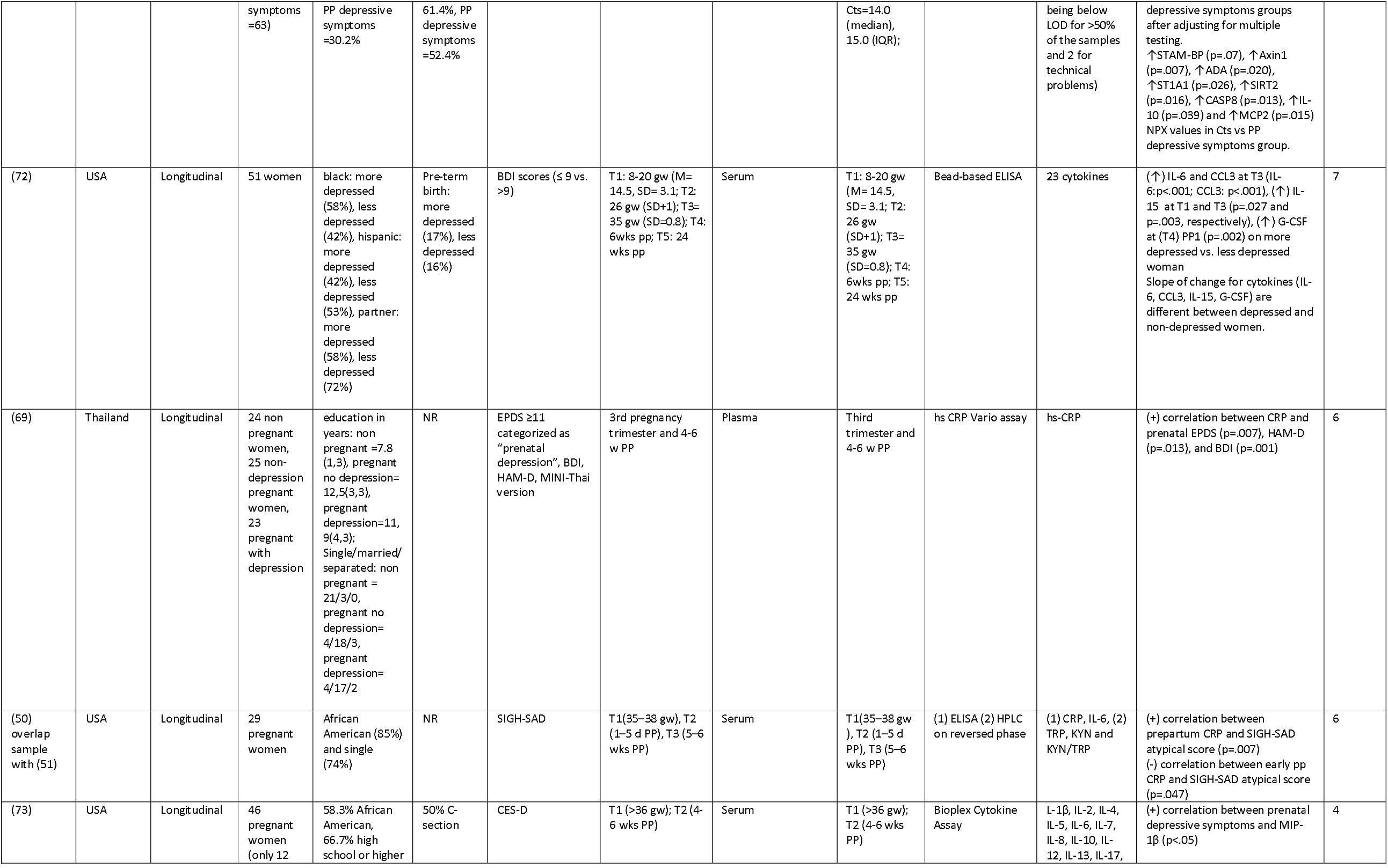
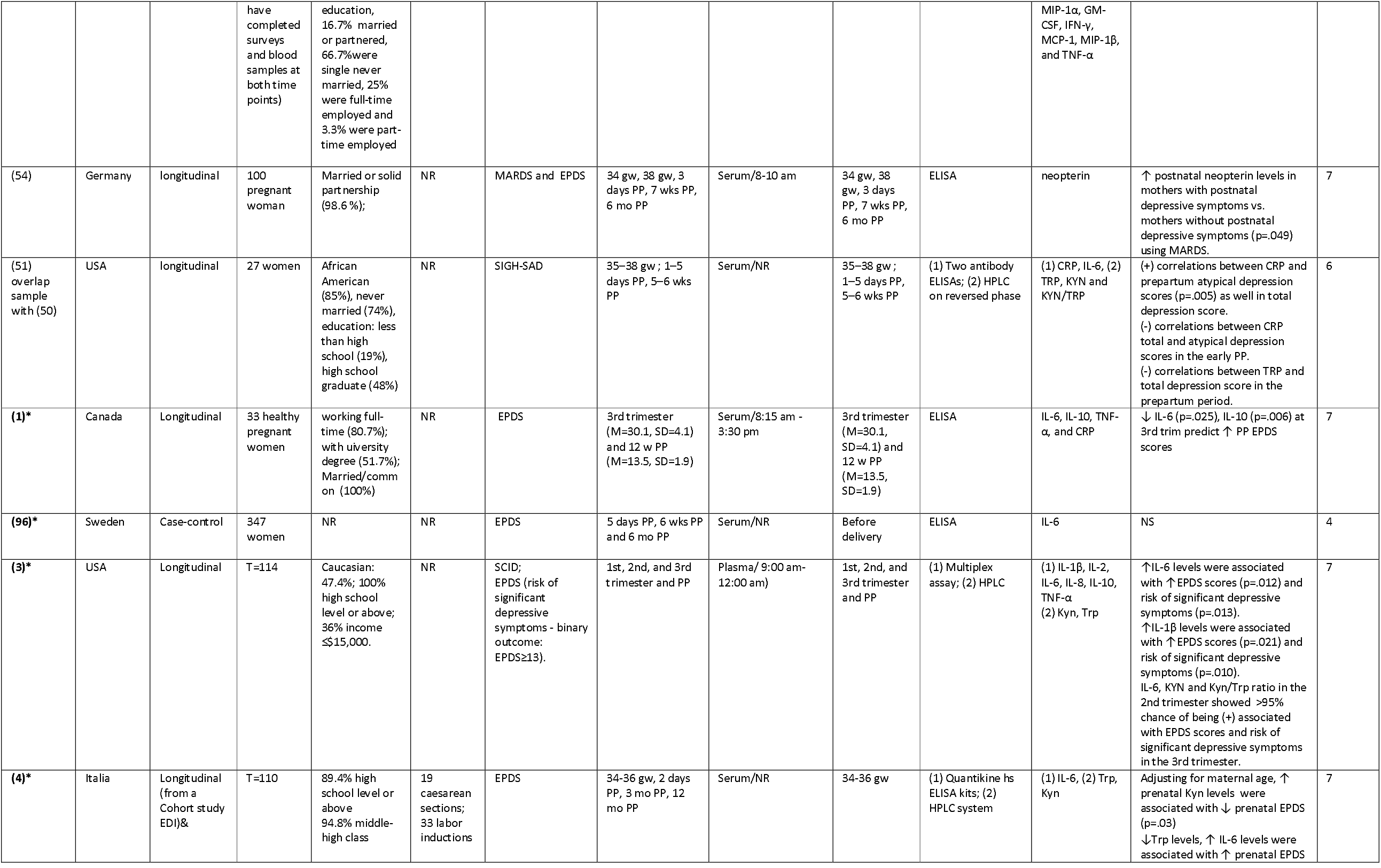
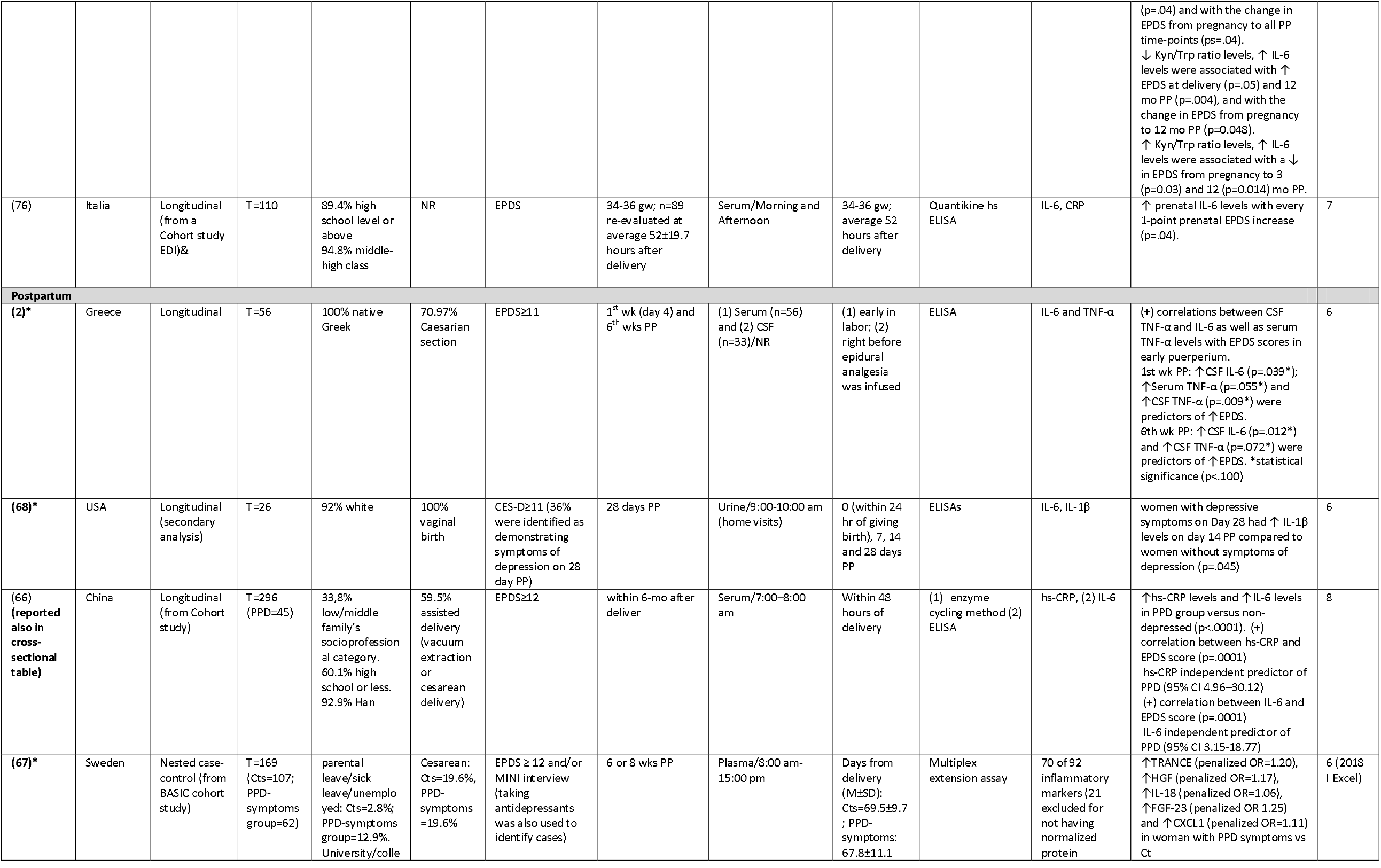
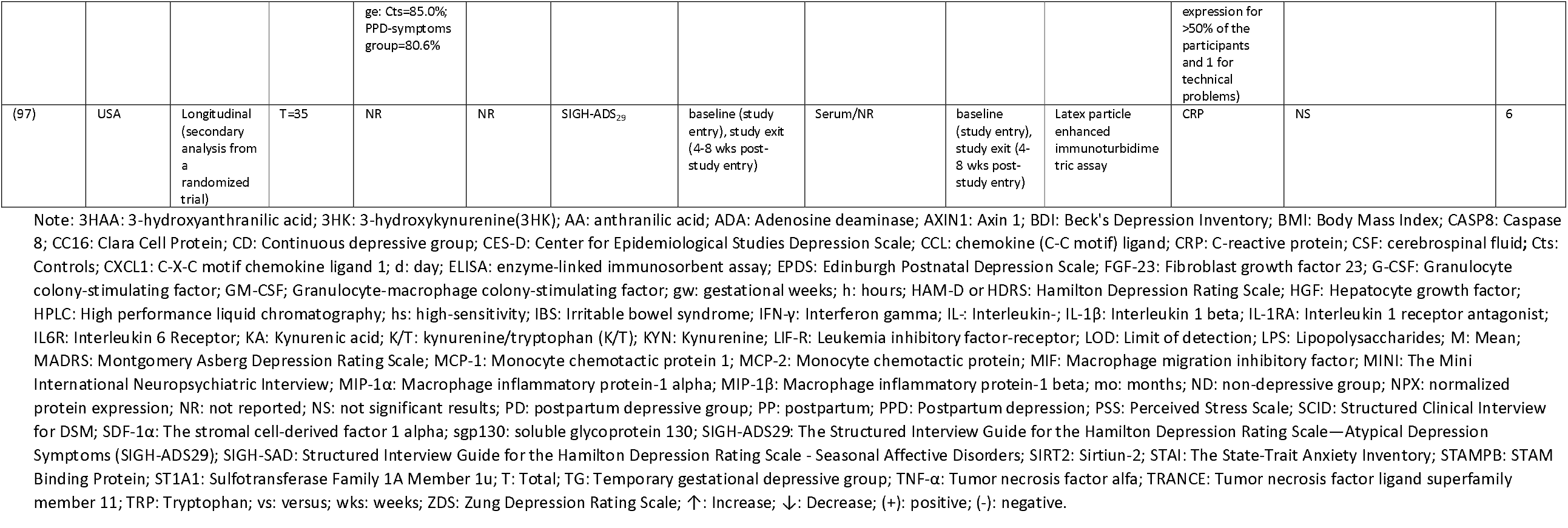
Longitudinal studies of inflammatory biomarkers and depression

### Quality assessment

Two researchers analyzed 56 articles for quality assessment. The inter-rater reliability between authors was analyzed calculating the agreement percentage (84%) and Cohen’s kappa (K=0.777), significance p-value < .001.

### Associations between inflammatory biomarkers and depression during pregnancy and during the postpartum period

#### Antenatal depression

Fourteen studies (4 case-control and 10 cross-sectional) exclusively assessed the association between depression and inflammatory biomarkers during pregnancy. Most of these studies were implemented during the 2nd and 3rd trimesters and used self-reported questionnaires to measure depressive symptoms / symptomatology (both using continuous scores and cut-off points) or depressive mood (33). Additional studies also included diagnosis of depression to define the depressed group (34-40).

Most of the studies found an association between several inflammatory biomarkers and depression status. In general, higher levels of mostly pro-inflammatory markers (namely, CRP, IFN-γ/IL-4 ratio, IL-1β, IL-1R, IL-5, IL-6, IL-8, IL-9, IL-12, IL-13, IL-23, IL-33 and TNF-α) were associated to higher depressive symptomatology or observed in depressed groups. For example, groups with higher depression symptoms demonstrated significantly higher circulating CRP levels (41) in comparison with those with lower depression symptom scores. Increased levels of CRP were also observed in a group with depression and trauma, in comparison to healthy pregnant volunteers (39).

Additionally, a positive correlation between CES-D scores and IL-1β (42) was found, as well as higher IL-1β in the CSF showing a significant association with increases odds of PND (37). Among other markers investigated, positive correlations were also found between IFN-γ /IL-4 ratio, IL-5, IL-9, IL-12 and IL-13 and EPDS scores (43). IL-1RA levels were significantly higher in women with high scores for depressive symptoms (CES-D>20) in comparison with women having scores less than 20 (44). Higher cerebrospinal fluid IL-1b, IL-23 and IL-33 concentrations at pre-cesarean time were significantly associated with increased odds of perinatal depression (37). Higher levels of TNF-α in depressed pregnant women were found when compared to controls in three studies (35, 38, 41). Nevertheless, these findings were not similar across all studies. Six studies had no significant results in (33, 34, 36, 40, 45, 46) (Table 1); one of them showing marginal p-values for higher CES-D scores as a predictor for higher levels of IL-6 and TNF-α (45).

In contrast with most of the studies in which a concrete small number of inflammatory proteins levels were determined, one study performed an immunoassay of a panel of 92 inflammatory proteins. Lower levels of 23 mostly anti-inflammatory proteins were found in women with antenatal depression (top three: TRAIL, CSF-1, CX3CL1) and women on SSRI treatment (top three: CSF-1, CEGF-A, IL-15RA) in comparison with controls (36) (Table 1).

In respect to markers representing the tryptophan kynurenine pathway, e.g. tryptophan (TRP) and/or kynurenine (KYN) and/or the ratio KYN/TRP were assessed in ten studies: two during pregnancy (41, 46); seven involving both pregnancy and postpartum (3, 4, 47-51); and one at postpartum (52). Findings point out some mixed results, considering the assessment time point considered and the statistical associations studied. In general, higher Kyn and Kyn/Trp ration was found in depressed group compared to controls, both during pregnancy (3, 47) and at postpartum (48). However, these results are not consistent, with no significant associations being found between KYN and KYN/TRP and depressive symptoms (51) or even describing the association in the opposite direction (4). Concerning TRP, findings showed no associations between TRP levels and prenatal depression (4, 51).

Among other markers investigated, most indicated a pro-inflammatory immune response in association with depressive symptoms, across the perinatal period; increase of LPS (53) and neopterin levels were found (54) whereas no differences were found for ERVWE1 levels (Human Endogenous Retrovirus WEnvC7-1 Envelope Protein) between women affected by PND and healthy controls (40).

#### Postpartum depression

The methodological diversity of the eleven studies assessing the association between inflammatory biomarkers and depression during postpartum is even larger than in the studies implemented during pregnancy. The time frame varied between one week and 12 months postpartum, but most studies were conducted between 4-12 weeks postpartum. Depressive symptoms or symptomatology was analyzed in four studies (#2, #48, #57, #83), depressive mood in three studies (#23, #45, #63), and diagnosis of depression in five studies. A diversity of inflammatory biomarkers assessed was also found, namely IFN-γ, IL-1α, IL-1β, IL-2, IL-4, IL-5, IL-6, IL-8, IL-10, IL12, IL-13, IL-17, IL-18, TGF-β2, TNF-α and other inflammation-related molecules.

Most of the studies found an association between inflammation and depression postpartum, except two studies (55, 56). Increased levels of IL-6 were associated with higher depressive symptomatology in two different studies (52, 57) (assessed with EPDS, SCID-5 and CES-D). Both (52, 57) were cross-sectional studies with the time point of assessment around 8 weeks postpartum. IL-8 levels were also in five studies and found to be positively associated with depression symptoms in 4 different studies all with a cross-sectional design (52, 57-59). The study of Fransson, E and colleagues (59), interestingly assessed how depression during late pregnancy affects inflammation around childbirth; this association was only in the group of women with premature delivery (representing 42 % of the total sample); the other 3 studies included later periods of assessment.

TNF-alpha, measured in four studies, was also shown to be significantly and positively associated with depression in one study (57), negatively associated with depressive symptoms when CpG-induced TNF-alpha was measured (60) and with no significant associations observed in the other two studies (52, 58). Specifically in the study from Christian et al. (57) the association between TNF-alpha and depressive mood (CES-D) was only found in a sample of African American women assessed between the 7 and 10 postpartum weeks.

Increased levels of other immunological mediators have also shown to be associated with depressive symptomatology, namely IL-2 (58), TGF-beta2 (61) and IgG (62). On the other hand, negative associations were described in 3 studies. Two found serum INF-gamma levels to be negatively associated with depressive symptomatology between the 4-6 weeks postpartum (63, 64). Also in a cross-sectional study, but congregating data from women participating at 12, 24 and 36 months postpartum, Gruenberg and collaborators have shown decreased induced-levels INF-gamma and other several cytokines, such as Il-8, TNF-alpha, IL-4, IL-5, IL-10 and IL-13, assessed from peripheral blood mononuclear cells associated with EPDS scores above 12 (60). Decreased levels of IL-2 have also been reported to be associated with increased risk for depressive symptomatology (52).

A lower ratio of KYN and of KYN/TRP ratio is observed during the postpartum period to that during pregnancy (47) and an increase in the K/T quotient over the postpartum period (48) was found in depressed group compared to controls. KYN levels and KYN/TRP ratio were found to be related with EPDS scores during postnatal period (47), and KYN/TRP ratio with the changes in EPDS from pregnancy to 12 months postpartum (4). Lastly, one study reported lower CC16 (considered anti-inflammatory) in women with postpartum depression (65).

#### Longitudinal studies, some including predictive approaches

Of the 31 longitudinal studies investigating the association between inflammatory biomarkers and depression over time, 4 were focused on the pregnancy period, five on the postpartum period and 22 across both pregnancy and postpartum. Various biomarkers were used for the investigations of association with depressive symptoms measured longitudinally (namely, 3HK and 3HAA, AA, CC16, CRP, GM-CSF, IFN-γ, IL-1β, IL-1RA, IL-2, IL-4, IL-5, IL-6, IL-6R, IL-7, IL-8, IL-10, IL-12, IL-13, IL-17, KA, KYN, leptin, LIF-R, LPS, MCP/CCL2, MCP-1, MIP-1α, MIP-1β, sgp130, TNF-α and TRP), although the most common findings regarded CRP, IL-6, TNF-alpha, KYN and TRP.

This study also demonstrated that IL-6 and TNF -alpha levels at birth were predictors of symptomatology at 1 and 6 weeks postpartum. This result was also corroborated by study (66), which also found IL-6 levels at delivery (within 48h) as an independent predictor of depressive symptoms assessed 6 months postpartum. Increased levels of other immunological mediators have shown to be associated with depressive symptomatology, namely CXCL1, FGF-23, HGF, IL-18, TRANCE (67), IL-1beta (68) and CRP (66).

Among the studies focusing C-reactive protein (CRP), three showed higher CRP levels in in association to prepartum depression and lower CRP levels in postpartum depression in the same two studies (50, 51) (sample overlap) and (69). The reports for TNF-α showed significant results in 4 studies. Although three of them showed lower TNF-α in PPD group (70) or in association with lower EPDS scores (24, 71), another study displays a higher TNF-α at 8-12 gestational weeks in depressed subjects vs. a healthy group, even when BMI, age and race were controlled for (53). Decreased levels of inflammatory markers were found in a depressive symptoms group vs. controls after adjusting for multiple testing, where women with PPD presented higher plasma levels for five inflammatory markers: CXCL1, FGF-23, HGF, IL-18 and TRANCE (25). The opposite was nevertheless true for the panel of 23 molecules considered in a study (72), where higher levels of inflammatory biomarkers were found in depressed group vs. controls; here, a different slope of change for cytokines (IL-6, CCL3, IL-15, G-CSF) was reported between depressed and non-depressed women. In turn, considering the 17 molecules assessed in the study (73) only MIP-1β showed a positive correlation with depressive symptoms.

Of the 22 longitudinal studies across pregnancy and the postpartum period, eight examined the potential predictive role of either inflammatory markers in the later depressive symptoms, or of depressive symptoms in predicting later inflammation profiles. One study showed higher depressive symptoms as a predictor for higher levels of IL-6 at midgestational period, as well as a significant association between increase in depressive symptoms from early to mid-gestation and IL-6 levels (74). In the same study, higher depressive symptoms at early or midgestational predict higher levels of CRP levels at midgestational period (74). Further, another study showed that lower IL-6 levels in the 3rd trimester predicted higher EPDS scores postpartum (1). In another study, family history of depression, third semester cortisol AUC, and third semester IL8/IL10 predicted symptoms of PPD (68). Moreover, increased IL-6 levels were found in depressed groups vs. controls at 24-28 gestational weeks (53), both before and after delivery (75). Additionally, higher IL-6 levels were associated with higher prenatal (3, 4) and postnatal EPDS scores (4), as well as with changes in EPDS across pregnancy (3, 4, 76). Lower prenatal Kyn levels were associated with greater depressive symptoms in late pregnancy, with prenatal Trp levels and Kyn/Trp ration moderating the association between IL-6 levels both antenatally and postpartum (4). One study found that cytokines and tryptophan metabolites predicted depression during pregnancy and that IL-1β and IL-6 levels were associated with severity of depression symptoms during pregnancy and postpartum (3). Centering in the longitudinal studies implemented during the postpartum period, the study conducted by Boufidou (2) showed that the TNF-alpha levels assessed in the CSF during labor significantly predicted increased depression symptoms at either 1st and 6th weeks postpartum, while serum CSF was only associated with the symptomatology at the 1st week postpartum.

## 4. Discussion

The present systematic review explored the association between a variety of inflammatory markers and depression, in different time points from pregnancy to postpartum period. Despite the large volume of available evidence stemming from 83 studies, a combined quantitative synthesis of all eligible studies was not feasible owing to the large variability in inflammatory markers assessed, the different study designs (cross-sectional, case-control and longitudinal studies), the different windows of exposure or outcome assessment, and the methods used for assessment for both depression and inflammatory markers.

Most studies assessed cross-sectional associations, while some few tried to assess the predictive potential of inflammatory markers for depression at later time-points, or of depressive symptoms to predict later inflammatory states. Overall, the findings of our systematic review lend support to the hypothesis that several inflammatory markers may be associated with peripartum depressive symptoms. The associations were somewhat different looking at pregnancy compared to the delivery time-point and postpartum, and mainly referred to increased levels of IL-6, IL-8, CRP and TNF-α among depressed. Evidence on the association of other inflammatory markers and PPD remains more inconclusive and replication studies are needed.

Inflammatory markers are a very broad family of heterogenous components, which have long been reported to play a significant role in the pathogenic pathways of several neurological and psychiatric diseases (77, 78). In addition, several molecules known to be activated in the inflammatory milieu or, on the other hand, having a role as inducers of an inflammatory response, are molecules of great interest on the molecular mechanisms of depression and others psychiatric diseases.

Congruent with previous studies on non-pregnant women with depressive symptoms (79-82), the most consistent finding of the present study was the significant association between elevated CRP levels and depressive symptoms during pregnancy. Pro-inflammatory markers, such as TNF-α, IL-1beta and IL-6 are released as response to stress or tissue damage, and they in turn induce the release of acute phase proteins, i.e. CRP, into the plasma. The molecular pathways through which these cytokines can impact on the development of depressive symptoms involves the dysregulation of neurotransmitter synaptic availability of monoamines such as serotonin, noradrenaline and dopamine, as well as the metabolism of various amino acids such as tyrosine, tryptophan, phenylalanine and glutamate (83). Tryptophan (TRP) metabolism plays an important role in the mechanisms associated with the gut-brain axis (84). Specifically, the kynurenine pathway (KP) is responsible for more than 90% of TRP catabolism throughout the body, with indoleamine 2,3-dioxygenase (IDO), the key metabolic enzyme, being activated in the inflammatory environment, leading to the downstream production of a variety of neuroactive compounds. The remainder of TRP is metabolized to serotonin and indole (85). In parallel, dysregulation of TRP metabolites such as serotonin, quinolinic acid (QUIN), and kynerunic acid (KA) has been linked to depressive behavior in animal models as well as in humans. Specifically, IL-1β and TNF-α may be responsible for the induction of p38 mitogen-activated protein kinase (MAPK), which in turn can increase the expression and function of serotonin reuptake pumps, resulting in decreased serotonin synaptic availability and subsequently in depressive-like behavior in experimental animal studies (86). Another biological mechanism that may underlie the association between inflammation and PPD onset includes the release of reactive oxygen or nitrogen species which in turn can decrease the availability of tetrahydrobiopterin (BH4), a key enzyme co-factor in monoamine synthesis (87).

The present review also highlights the large heterogeneity of results regarding the role of different inflammatory markers in depression during different time periods. This can be at least partly explained by the window of exposure; compared to delivery and postpartum, pregnancy is a period of large HPA-axis and sex-steroid hormone changes, which may explain the robust findings of the identified studies focusing on inflammatory markers during pregnancy (88). An earlier clinical trial (75) reported that the levels of IL-6 and its receptor (IL-6R) were significantly higher during early pregnancy than before delivery, and women who developed depressive symptoms in the early puerperium had significantly higher serum IL-6 and IL-6R concentrations than those without. However, the different timing of sampling, even the week of sampling, the different study designs (cross-sectional versus longitudinal, population-based vs. case-control, etc.), as well as the different power of each study and diversity of ethnicities among the study populations can also account for eventual differences in the results.

An important finding of the present review was the inclusion of longitudinal studies which assessed the role of inflammatory markers as predictive markers in depressive symptoms onset. In particular, six studies assessed biomarkers at some time during pregnancy in relation to depression during postpartum, showing a potential predictive role of TNF-a and IL-6 in the diagnosis of PPD (1, 3, 24, 70, 71, 75). Moreover, three studies with longitudinal design explored the predictive role of inflammatory markers assessed during delivery in relation to depression during postpartum (2, 66, 67). These results are congruent with a recent review reporting the predictive value of proinflammatory cytokines in the diagnosis of PPD during postpartum (89). In general, although longitudinal studies suggest that the relationship between depression and inflammation is characterized by complex bidirectional associations, existing, prospective, longitudinal research designs are still poorly equipped to investigate the dynamic interplay of depression and inflammation that unfolds over a relatively short time period (90).

### Critical appraisal: strengths and limitations

The present study acknowledges that the systematic review of PPD epidemiology, especially on the role of inflammatory markers in PPD onset, is a rather challenging field of research mainly due to the large heterogeneity of available evidence and several inherent limitations of the individual studies. First, the definition of exposure and outcome among inflammation and depression is not always straight-forward. Further, the large heterogeneity in assessment of the inflammatory markers is perhaps the most important methodological limitation of the studies. Markers of inflammation are a heterogeneous group of very different active components. The studies have often focused on different molecules and have even used different methods for their identification and quantification. In addition, the identified studies assessed the role of inflammatory markers in different time points, namely during pregnancy, delivery and postpartum, thus not readily allowing a quantitative synthesis of the results in the context of a meta-analysis. Further, even depression was assessed differently, with some studies using self-reports measures that often capture perinatal distress and not depression in particular, while other used clinical instruments used in psychiatric settings to set a diagnosis of major depression. This might account for some of the inconsistencies in results between the studies. Among other limitations, we excluded from this systematic review results of inflammatory markers related to quantification of immune cells as well as physical properties such as erythrocyte sedimentation rate (ESR). Moreover, mRNA and epigenetic studies were also excluded, as this work focused on protein level markers. In addition, it would be essential to account for confounding from potential co-exposure to multiple markers or even other molecules and hormones, in order to delineate unbiased associations, but this has not been possible in the overwhelming majority of evidence assessed herein.

Nonetheless, beyond these limitations, the present study followed a strict pre-registered protocol and systematically reviewed all available evidence regarding the association between different inflammatory markers at the protein level and peripartum depressive symptoms. In trying to synthesize the available evidence, we also grouped the results by window of exposure and type of study, and identified studies where inflammatory markers were tested for their predictive potential.

## Conclusions and practical clinical implications

The present systematic review summarized the current evidence on the association of inflammatory markers and depressive symptoms during the peripartum period. Beyond potential limitations and biases, the findings of the present work provide evidence of increased pro-inflammatory markers among women with depressive symptoms. Further evidence from sufficiently powered studies that apply robust and preferably the same methods for assessment of inflammatory markers and PND symptoms, at specific and homogeneous time periods is needed; this could further enhance our understanding of the pathophysiology of PND with mechanistic studies, and shed light into potentially relevant molecular pathways that underlie these associations, in the hope of new possible treatments. Further work could point to the practical application of a panel of such biomarkers for the early identification of women at risk of developing PPD in order to use preventive interventions.

## Data Availability

All relevant data are within the manuscript and its Supporting Information files.

## Acknowledgements

The authors would like to thank Maria Karalexi, Pietro Gambadauro, Georgios Schoretsanitis and Andrea Hess Engström for help with appraisal of results, and Mariana Saraiva for support on title and abstract screening.

Authors declare no conflicts of interest.

## Supporting information

**S1 Table. Search terms**.

**S2 Table. Studies reporting results between depression and inflammation as secondary data**.

