## Supplementary material for "Inflammatory biomarkers and perinatal depression: a systematic review": S1 Table

| **Pubmed** | **Psycinfo** | **Web of science core collection** |
| --- | --- | --- |
| (depress* OR “Depression”[Mesh] OR “Depression, Postpartum”[mesh] OR “Depressive disorder”[mesh]) AND (perinatal OR peripartum OR postpartum OR postnatal OR antepartum OR antenatal OR childbirth OR pregnan* OR “Peripartum Period”[mesh] OR “Postpartum Period”[mesh]) AND (immun* OR inflamm* OR cytokine* OR interleukin* OR chemokine* OR “c-reactive protein” OR “tumor necrosis factor*” OR interferon* OR “Cytokines”[mesh] OR “Interleukins”[mesh] OR “Chemokines”[mesh] OR "C-Reactive Protein"[mesh] OR "Tumor Necrosis Factor-alpha"[mesh] OR “Interferons”[mesh] OR “Inflammation”[mesh] OR “Immune System”[mesh])  Filters:  journal article  Humans  language (English, Portuguese, spanish, French) | depress*.mp. or depression.sh. or “postpartum depression”.sh. or “depressive disorder*”.mp. and (perinatal or peripartum or postpartum or postnatal or antepartum or antenatal or childbirth or pregnan*).mp. or “perinatal period”.sh. or “postnatal period”.sh. and (immun* or inflamm* or cytokine* or interleukin* or chemokine* or “c-reactive protein” or “tumor necrosis factor*” or interferon*).mp. or “Cytokines”.sh. or “Interleukins”.sh. or "tumor Necrosis Factor”.sh. or “Interferons”.sh. or “Inflammation”.sh. or “Immune System”.sh.  Filters:  All journals  Human  language (English, Portuguese, spanish, French) | TS=(depress* OR postpartum depression OR depressive disorder) AND TS=(perinatal OR peripartum OR postpartum OR postnatal OR antepartum OR antenatal OR childbirth OR pregnan*) AND TS=(immun* OR inflamm* OR cytokine* OR interleukin* OR chemokine* OR C-reactive protein OR tumor necrosis factor* OR interferon* OR immune system)  Filters:  TIPOS DE DOCUMENTO: (Article)  IDIOMA: (English OR French OR Portuguese OR Spanish) |
