## Supplementary material for "Inflammatory biomarkers and perinatal depression: a systematic review": S2 Table

**Supplementary Table :** Studies reporting results between depression and inflammation as secondary data.

| **Reference** | **Country** | **Study design** | **Number of subjects** | **Socio-economic status/etnicity** | **Delivery mode** | **Assesment of depression** | | **Inflammatory protein markers** | | | | **Results** |
| --- | --- | --- | --- | --- | --- | --- | --- | --- | --- | --- | --- | --- |
|  |  |  |  |  |  | **Instruments** | **Timepoint(s)** | **Biological fluid/hour of collection** | **Timepoint(s)** | **Dosage assesment technique** | **Inflammatory markers** |  |
| (Groer et al., 2018) | USA | Cross-sectional | 374 pregnant women TPO negative | 19.6%: less than college education.  16%: incomes less than US$25,000/year. 45.6%: Caucasian | NR | POMS-D | 19.8±2.8 gw (range=15.2–25.6) | (1) serum and plasma; (2) serum; (3) serum | 19.8±2.8 gw (range=15.2–25.6) | (1) HPLC; (2) ELISA; (3) Griess method | (1) TRP and KYN; KYN/TRP ratio; (2) Neopterin; (3) Nitrite level | ↓ TRP in women with POMS-D scores >20 compared to lower POMS-D scores (p=.017).  (+) correlation between POMS-D scores and nitrite levels (p=.04). |
| (Osborne et al., 2018) | UK | Longitudinal | 106 pregnant women (49 cases with MDD in pregnancy and 57 healthy controls) | Higher deprivation in cases compared with controls (IMD) | NR | SCID I–CV and BDI | SCID I–CV: 25 gw (baseline). BDI: 25 and 32 gw; 6-days, 2- and 12-mo PN. | Serum | Median = 37 gw (range 23.9-34.9 gw) | (1) ELISA; (2) cytokine chip array kit | (1) hs-CRP, (2) IL-1β, IL-2, Il-6, IL-8, IL-10, TNFα, VEGF, EGF, MCP-1. IL-1α, IL-4 and INF-γ, > 50% bellow detection level | ↑ IL-6 (p=.031), ↑ IL-10 (p=.043), ↑ TNFα(p=.003) and ↑ VEGF (p=.008) in cases compared to controls (differences remained significant after adjusting for IMD; Only TNFα remained significant after adjusting for multiple comparisons) |
| (Raw et al., 2014) | USA | Case-control | 63 healthy pregnant women (21 depressed and 42 matched Cts) | 71.4% African American  39% (MDD group) and 46% (Ct group) <10000$ income | NR | EPDS (EPDS >=15 MDD cases) | Ct: 10.6 ± 3.1 gw MDD: 11.2 ± 2.9 gw | Plasma | 1st trim (6-16 gw), 2nd trim (16-26 gw), 3rd trim (30-37 gw) and at admission for delivery | ELISA | hs-CRP | ↑ CRP in MDD vs CT group at the 1st trim (p=.027) |
| (Nazzari et al., 2019) | Italy | Longitudinal | 104 pregnant women | 97.1% Italian, 89.4% high school diploma, 94.8% middle-high class | 82.7% vaginal delivery and 98.1% full term | EPDS | 30–33 gw (31.45 ± 1.40) | Serum | 34–36 gw (34.76 ± 1.12). | hsELISA | IL-6, CRP | (+) correlation between EPDS and IL-6 (p<.05) |
| (Bjelanović et al., 2015) | Bosnia and Herzegovina | Cross-sectional | 180 pregnant women | NR | NR | Symptoms of depression - ICD-10; Depression symptomatology Beck self depression scale; Depression - SCL90-R . | third trimester of pregnancy - 48h after delivery | Blood | third trimester of pregnancy - 48h after delivery | Clinical analysis using turbidimetry method | CRP | (+) correlation between CRP and depression (p=.001), number of depressive symptoms (p=.001) and Beck scores (p<.001). |
| (Okun et al., 2013) | USA | Longitudinal (from a Prospective study) | T=168 (Ct=136, depressed=32) | 16% high school or less education; 80.2% white race | NR | SCI-DSM-IV | 20 gw | Plasma/9:30am-1:30 pm | 20 and 30 gw | ELISA | IL-6, IL-8, IFN-γ and TNF-α | NS |
| (Pan et al., 2018) | China | Case- control | 56 women with PPD and 27 matched controls | NR | NR | EPDS score ≥ 10 | 6 wks after delivery | Serum | 6 wks after delivery | ELISA | IL-1β, CXCL2, CXCL3 | ↑ IL-1β (p<.001), CXCL2 (p<.0001) and CXCL3 (p<.001) in EPDS low vs. controls (EPDS = 0);  ↑ IL-1β, CXCL2 and CXCL3 (p<.001) in EPDS high vs. controls (EPDS = 0) (p<.0001);  ↑ IL-1β, CXCL2 and CXCL3 (p<.001) in EPDS high vs. EPDS low (p<.0001) |
| (Aparicio et al., 2020) | The Netherlands | Longitudinal | T=51 | 16% educational background middle job trainning | NR | EPDS | 6 wks PP | Human milk/08:36  ±02:48 am | 2, 6, and 12 wks PP | (1) Multiplex immunoassay; (2) ELISA; (3) Isotyping  Assay  kit | (1) IL1β, IL6, IL12, IFNγ, TNFα, IL2, IL4, IL10, IL13, IL17, IL8, Groα, MCP1, MIP1β, IL5, IL7, GCSF, GMCSF, TGFβ2; (2) EGF; (3) IgA, total IgG, IgM, | (+) correlation between IL-7 and depressive symptoms (p=.040) at 6 wks PP. NR for 2 and 12 wks PP. |
| (Robertson Blackmore et al., 2016) | USA | Longitudinal cohort study | 171 women | 47.4% African American, 20.5% lifetime exposure to intimate partner violence | 34.2%Cesarean section, 13% Premature birth | (1) SCID; (2) EPDS | (1) 18 and 32 gw (+\- 1w), and 6wks PP and 6 mo PP  (2) 32 gw | Serum | 18 and 32 gw (+\- 1w), and 6w PP and 6 mo P | hsELISA | IL-6, TNF-⍺ | NS (depression as covariate in multivariable analysis) |
| (Accortt et al., 2016) | USA | Longitudinal | 91 pregnant women | 100% African American  Married (25%), Employed (63%), ≥High school diploma (58%)  Annual income $36,623±34,609 | NR | EPDS | 4-6 wks PP (M=4.5±1.8) | Serum | 13-28 gw (M=21.3±3.8) | (1) enzyme immunoassay (BioCheck); (2) Bio-Plex Pro | (1) hs-CRP; (2) IL-1β, IL-6, IL-10, and TNF-α | NS |
| (Accortt et al., 2016) | USA | Longitudinal | 68 pregnant women | 76.7% non-Hispanic Caucasian | NR | CES-D | T1 (24 gw), T2 (37 gw), T3 (6 mo PP) | Plasma | 37 gw | hsELISA | IL-6, TNF-⍺, MCP-1, IL-10 | Maternal prenatal depressive symptoms were associated with maternal inflammation at the 3rd trim (IL-6, TNF-⍺, MCP-1) (p<.001)  (+) correlation between prenatal CES-D and 3rd trim IL-6 and TNF-⍺ (p<.05), MCP-1 (NS) |
| (Nazzari et al., 2020) | Italy | Longitudinal | 104 pregnant women | 97.2% Italian  87.5% middle-upper class | 81.7% vaginal delivery | EPDS | 34-36 gw (M=34.76±1.12) and 12 wks PP (M=11.96±1.85) | Serum | 34-36 gw (M=34.76, SD=1.12) | hsELISA | CRP and IL-6 | NS correlations between prenatal EPDS and prenatal IL-6 and CRP |
| (Finy & Christian, 2018) | USA | Cross-sectional | 214 pregnant women | 65.9% White  22.4% < $ 15,000  31.8% graduate school | NR | CES-D | 5-31 gw (M=17.7±7) | Serum | 5-31 gw (M=17.7±7) | (1) solid-phase chemiluminescence immunometric assay; (2) electrochemiluminescence | (1) CRP, (2) IL-6 | NS correlation between CES-D and CRP; (+) correlations between CES-D and IL-6 (p<.01) |
| (Groër et al., 2011) | USA | cross-sectional | 414 pregnant women | NR | NR | POMS-D | 16-25 gw | Plasma | 16-25 gw | (1) Luminex technology ; (2) high-performance liquid chromatography; (3) ELISA | (1) IFN-γ, IL-10, TNF-α; (2) TRP, KYN, TRP/KYN ratio; (3) Neopterine | Only reported results for TRP: ↓TRP in depressed group (POMS-D>20) vs nondepressed (p=.02) |
| (Morgan et al., 2020) | USA | longitudinal | 100 mother children dyads | Household income: $66,363± 63,027 | NR | EPDS, CES-D | preconception (5.93 mo ± 6.07), 2nd trim (20.25±4.54 gw), 3rd trim (32.85±3.26 gw) | Whole blood spots on Guthrie paper NR? | preconception (5.93 mo ± 6.07), 2nd trimester (20.25±4.54 gw), 3rd trimester (32.85±3.26 gw) | NR | hs-CRP | NS |
| (Ruyak et al., 2016) | USA | longitudinal | 111 pregnant women | 82.9% white | 100%uncomplicated vaginal birth | EPDS | 32- 36 gw; 4 wks PP | plasma | 32- 36 gw; 4 wks PP | Proinflammatory Ultra-Sensitive assay | IL-6, TNF-α | NS standardized coefficient for the path from inflammation to symptoms of depression at 4 wks PP (mediation model) |
| (Giurgescu et al., 2016) | USA | cross-sectional | 96 pregnant African American women | 50% annual household income < $10 000  51% unemployed some college (39.6%) | NR | CES-D | 15-26 gw (19.7 ± 2.5 gw) | plasma | 15-26 gw (19.7 ± 2.5 gw) | Multiplex bead immunoarrays | L-1β, IL-2, IL-4, IL-6, IL-8 and IL-1 | NS only reported for IL-4 and IL-6 in multiple linear regression models. |
| (Zhu et al., 2020) | USA | Case-control study | 74 pregnant women | 40.5% African American  52.7% Single/divorced | NR | PHQ-9 | 24-36 gw (M=32.8±3.5) | serum | 24-36 gw (M=32.8± 3.5) | Ultrasensitive ELISA | CRP, sIL-6R, IL-6 | (+) correlations between PHQ-9 scores and morning CRP (p<0.05) |
| (Groer et al., 2020) | USA | cross-sectional pilot study | 97 women | 72% unemployed, 54% college or postgraduate education preparation; 55% with annual income >$40,000, 28% single/divorced  76% Caucasian | NR | POMS-D | 2 -4 mo PP | ex-vivo cultures fluid | 2 -4 mo PP | ELISA | IL-6 | NS for POMS-D (in a sub-sample of 59 women) |
| (Christian et al., 2013) | USA | Cross-sectional | T=101 | 52.5% High school or less; 63.4% household income <$15,000; 54.5% African-American | NR | CES-D | M range 23.2-24.6 gw | Serum/8:00am–4:00pm | Mean range 23.2-24.6 gw | (1) Multiplex assay, (2) hs-ELISA | (1) IL-6, TNF-α, IL-1β, (2) CRP, MIF | (+) correlation between depressive symptoms and TNF-α (p<.05)  (-) correlation between depressive symptoms and IL-1β (p<.05) |
| (Ahn & Corwin, 2015) | USA | Longitudinal | 119 women | 82% Caucasian (68–82%), 96% married | 100% vaginal birth | EPDS | 32-36 gw, 1 wk PP, 2wks PP, 1 mo PP, 2 mo PP, 3 mo PP, 6 mo PP | plasma | 32-36 gw, 1 wk PP, 2 wks PP, 1 mo PP, 2 mo PP, 3 mo PP, 6 mo PP | Human Pro-inflammatory Ultra-Sensitive assay and quantitative multiplex array technology | IL-6, IL-1β, IL-8, IFN-γ, TNF-α, IL-10 | NS correlations between EPDS scores and IL-6 at 6 mo PP |
| (Albacar et al., 2010) | Spain | Case-control | 1053 women | Depressed/non depressed: 39.1%/30.4% primary school; 16.1%/13.5% unemployed; 2.3%/1.9% single | NR | EPDS, Diagnostic Interview for Genetics Studies (DIGS) applied to all probable cases of major depression | 24-48 PP, 8 wks PP, 32 wks PP | plasma | 2 d PP | Ultrasensitive turbidimetric immunoassay | CRP | NS differences between depressed and non-depressed participants |
| (Hunter et al., 2021) | USA | Prospective study | T = 201 pregnant women | Maternal education: males fetuses=13.7±3.0 and females fetuses=13.4±3.1 | NR | CES-D | 16 gw | Plasma | 16 gw | 1) Beckman–Coulter high sensitivity assay; 2) R&D Systems high sensitivity assays | 1) CRP; 2) TNFα, IL-6 and IL-8 | (+) correlation between CRP and CES-D scores (p=.03) |
| (McCormack et al., 2021) | USA | Longitudinal study | T=186 | 14,5% Income ≤$15,000; 69.4% Hispanic ethnicity | NR | interviewer-administered HAM-D | T2 (24-27 gw) and T3 (34-37 gw) | Plasma/(8:00 am-6:00pm) | T2 (24-27 gw) and T3 (34-37 gw) | hs ELISA | IL-6 | (+) correlation between T3 IL-6 and T3 HAM-D (p=.05) |
| (Freedman et al., 2021) | USA | Cohort | T=162 (CESD rating≥16: n=61; CESD rating<16: n=101) | 57% native American in CESD≥16 group and 67% European American in CESD<16 group. | NR | CESD | 16 gw (range, 15–17 wks) | Plasma/midmorning | 16 gw (range, 15–17 wks) | hs ELISA | CRP | ↑ CRP levels in CESD≥16 vs CESD<16 groups only with male fetuses (p=.016) |
| (Nagayasu et al., 2021) | Japan | Longitudinal | T=80 pregnant women (EPDS≤7: n=67 and EPDS>7: n=13 groups) | NR | Cesarean section: EPDS≤7=22.2%; EPDS>7=26.7% | EPDS | 1 mo PP | Serum/morning | 1^st^ and 2^nd^ trimesters,and 3–5 days PP | ELISA | IL-6 | NS |
| (Gillespie et al., 2021) | USA | Secondary analysis from a prospective cohort | T=93 (full analytical sample) | 100% non-Hispanic Black American women; | NR | CES-D (≥16 minor perinatal depression; ≥24 major perinatal depression) | Early third trimester  of pregnancy: 30 gw and 2.3 days of pregnancy (±1week 5.91 days) | Plasma/13:00 hours±93.6 min | Early third trimester  of pregnancy: 30 gw and 2.3 days of pregnancy (±1 week 5.91 days) | Multiplex assay | IL-6, IL-8, TNF-α and IL-1β | NS |

Note: BDI: Beck's Depression Inventory; CES-D: Center for Epidemiological Studies Depression Scale; CRP: C-reactive protein; Cts: Controls; CXCL: C‐X‐C motif chemokine ligand; d: day; EGF: Epidermal growth factor; ELISA: enzyme-linked immunosorbent assay; EPDS: Edinburgh Postnatal Depression Scale; G-CSF: Granulocyte colony-stimulating factor; GM-CSF; Granulocyte-macrophage colony-stimulating factor; Groα: growth-related oncogene alpha; gw: gestational weeks; HAM-D: Hamilton Depression Rating Scale; HPLC: High performance liquid chromatography; hs: high-sensitivity; ICD-10: the tenth revision of the International Classification of Diseases and Causes of Death; IFN-γ: Interferon gamma; Ig: Immunoglobulins; IL-: Interleukin-; IL-1α: Interleukin 1 alpha; IL-1β: Interleukin 1 beta; sIL-6R: Interleukin 6 soluble Receptor; IMD: the Index of Multiple Deprivation; KYN: Kynurenine; M: Mean; MCP-1: Monocyte chemotactic protein 1; MDD: Major Depressive Disorder; MIF: Macrophage migration inhibitory factor; MIP-1β: Macrophage inflammatory protein-1 beta; mo: months; NR: not reported; NS: not significant results; PHQ-9: The Patient Health Questionnaire; PN: Postnatal; POMS-D: Profile of Mood States-depression–dejection scale; PP: postpartum; PPD: Postpartum depression; SCI-DSM-IV: SCID: Structured Clinical Interview for DSM; SCL90-R: standardized psychometric Questionnaire; T: Total; TGF-β2: Transforming growth factor-beta 2; TNF-α: Tumor necrosis factor alfa; TPO: Thyroid peroxidase; Trim: Trimester; TRP: Tryptophan; VEGF*: V*ascular Endothelial Growth Factor; vs: versus; wks: weeks; ↑: Increase; ↓: Decrease; (+): positive; (-): negative.
